# Comprehensive genome analysis of 6,000 USA SARS-CoV-2 isolates reveals haplotype signatures and localized transmission patterns by state and by country

**DOI:** 10.1101/2020.05.23.20110452

**Authors:** Lishuang Shen, Jennifer Dien Bard, Jaclyn A. Biegel, Alexander R. Judkins, Xiaowu Gai

## Abstract

Genomic analysis of SARS-CoV-2 sequences is crucial in determining the effectiveness of prudent safer at home measures in the United States (US). By haplotype analysis of 6,356 US isolates, we identified a pattern of strongly localized outbreaks at the city-, state-, and country-levels, and temporal transmissions. This points to the effectiveness of existing travel restriction policies and public health measures in controlling the transmission of SARS-CoV-2.

---

SARS-CoV-2 is a positive-sense single-stranded RNA virus (Wu et al, 2020; Zhu et al., 2020). The collection of variants in a viral genome is referred to as the haplotype. New haplotypes arise from sequential acquisition of new variants in the viral genome. A haplotype, more than individual variants, serves as the distinct signature of a viral isolate and can be used effectively to trace the lineage, determine the ancestral origin of the infection, and to understand the community spread pattern during the COVID-19 pandemic. Viral genomes and demographic meta-data of 6,356 SARS-CoV-2 isolates within the US (as of May 20, 2020) were extracted from GISAID (https://www.gisaid.org/), GenBank (https://www.ncbi.nlm.nih.gov/genbank/sars-cov-2-seqs/), and COVID-19 patients and staff at the Children’s Hospital Los Angeles (CHLA). Variants, haplotype, geographic location at diagnosis and documented exposure for the patients were analyzed (Shen et al. 2020). A total of 921 variants and 264 haplotypes were detected in at least three US isolates independently, that were deemed reliable and less likely to be sequencing artifacts (**Table S1**). The four most common mutations (241-C-T, 3037-C-T, 14408-C-T, 23403-A-G) were each present in about 65%-67% of US isolates. In total, these 921 variants included 487 missense, 348 synonymous, 66 intergenic, 4 in-frame deletions, 5 stop gained/lost, and several other noncoding variants (**Table S2**).

Cross-stratification by geolocation identified city-, state- and country-specific haplotypes. There were 849 isolates from the 77 US-specific haplotypes represented by at least five isolates, which together accounted for 13.3% of the US isolates, and 29.2% of the 264 total haplotypes found in the US isolates. In addition to the 77 purely US-specific haplotypes, there were an additional four large haplotypes that were mostly North America-specific, with a total of 434 isolates where 431 isolates (99.3%) were from the US (425) and Canada (6) (**Table S3**). Isolates from these four large haplotypes were geographically dispersed across the nation. Of note, all 66 US isolates belonging to haplotype (241-C-T, 1059-C-T, 3037-C-T, 11916-C-T, 14408-C-T, 18998-C-T, 23403-A-G, 25563-G-T, 29540-G-A) were from the COVID-19 epicenter in New York and neighboring states. Comprehensive phylogenetic analysis of the US-specific isolates, along with 2,000 randomly selected non-US isolates, revealed that the isolates fell exclusively in some major clades and were completely absent in the remaining clades (**Figure 1**). The mean number of isolates represented by each USA-specific haplotype was 11.4±12.9 (range: 5-91) (**Table S3**). Of note, 58 of the purely US-specific and the four nearly US-specific haplotypes consisted of 715 US isolates in total (52%) that all had the globally dominant 23403-A-G (D614G) mutation (Korber et al., 2020). The 8782-C-T (orf1ab, synonymous) and 28144-T-C (orf8:p.Leu84Ser) variants were mutually exclusive with D614G, and co-occurred in 25 haplotypes that accounted for a total of 551 US isolates.

**Figure 1.**
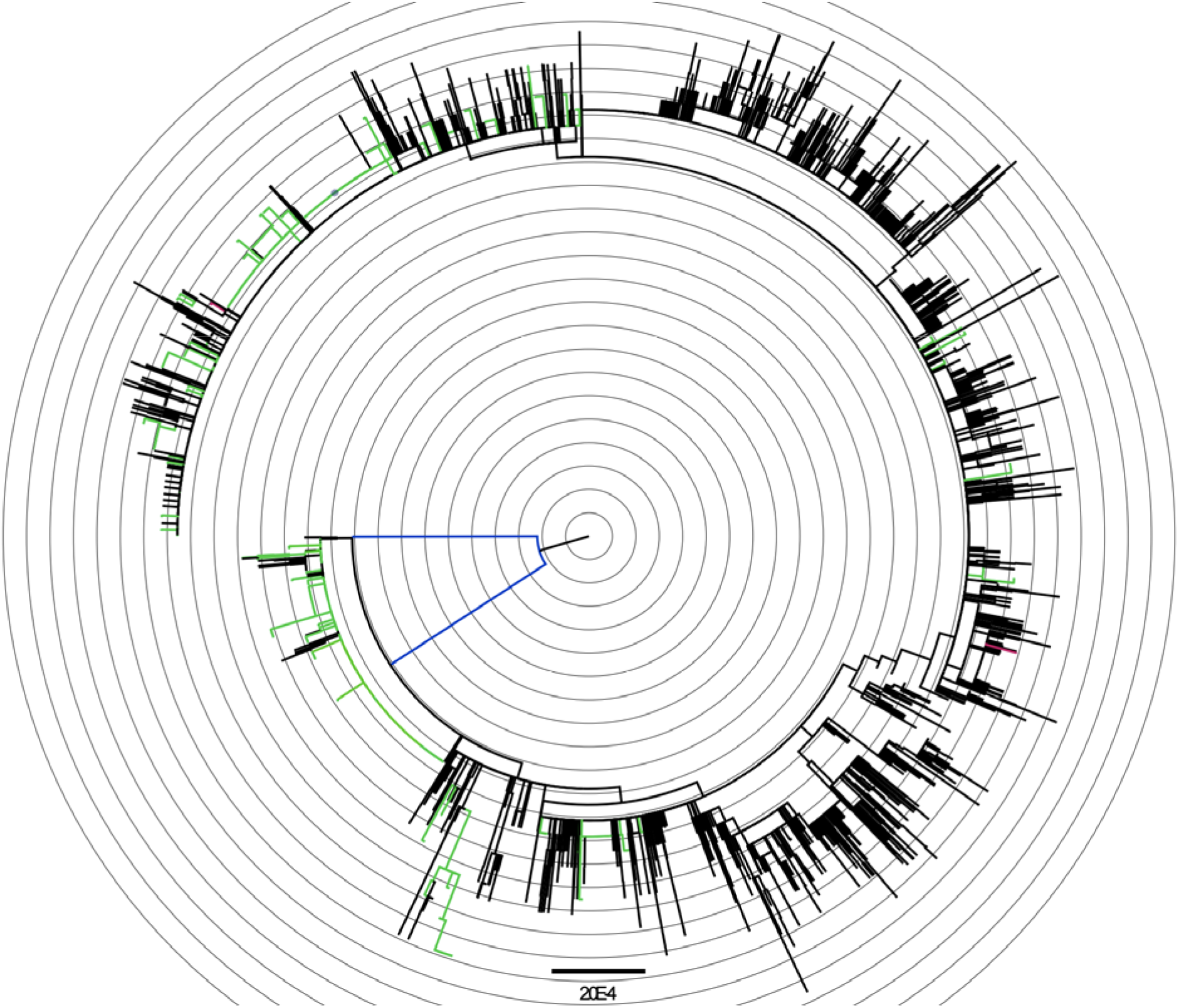
Maximum likelihood phylogenetic tree of isolates carrying US-specific haplotypes and 2,000 randomly selected non-US isolates. Green: isolates with US-specific haplotypes; Red: Children’s Hospital Los Angeles (CHLA) isolates with US-specific haplotypes; Black: Non-US isolates. Blue: Outgroup (MN996532|EPI_ISL_402131 bat/Yunnan/RaTG13/2013) and reference NC_045512 (MN908947|EPI_ISL_402125). 1,107 isolates from US-specific haplotypes were included in the phylogenetic analysis. Non-US isolates belonging to any haplotypes were randomly sampled at about 10% to reach 2000 isolates. Each branch in the phylogenetic tree may represent a group of isolates. The tree is rooted at the outgroup MN996532.

State-specific haplotypes were identified for 12 states with a total of 613 isolates out of 6,356 US isolates (9.6%). Seven US-specific haplotypes were almost exclusively found in isolates from a single state, while in many cases the exclusivity-violating isolates were from neighboring states. Washington state (28 haplotypes & 382 isolates) had the most private haplotypes, followed by California (9 haplotypes & 63 isolates) and Utah (6 haplotypes & 48 isolates). This may reflect early safer-at-home orders implemented in the states of Washington and California where the earliest COVID-19 cases were reported. The number of haplotypes increased over time as new variants were continuously acquired, but the newly emerged haplotypes were confined within these states. Two of the California specific haplotypes are notable. The 9-isolate 491-G-A,14940-A-G haplotype group and its single-marker ancestral haplotype (14940-A-G with 4 isolates) were exclusively present in California between March 31, 2020 and May 1, 2020. 491-G-A is a missense variant, p.Ala76Thr, in the orf1ab gene. The 15-isolate 25692-C-T haplotype group is similarly interesting in the sense that these haplotypes are relatively “ancestral” with only 1-base difference from the reference isolate genome (NC_045512.2), but they were recently seen in late April, after the inception of the safer at home policy in California. This is in contrast with the dominant haplotypes in the US that were more distant descendants of NC_045512.2, with at least 3 and frequently more than 10 variants compared to the reference. This is suggestive of containment of early infections in California and limited spread to other states, likely again because of the early response to the pandemic from the state of California.

On the national level, one major haplotype (8782-C-T, 17747-C-T, 17858-A-G, 18060-C-T, 28144-T-C) had 317 member isolates, where 315 (99.4%) were from the US (311, 98.0%) and neighboring Canada locations (4, 1.3%). With the exception of two isolates from Australia, there were no isolates from outside North America. It is noted that this haplotype lacks the dominant D614G mutation prevalent in Europe. The first reported USA COVID-19 case in mid-January, haplotype (3 variants 8782-C-T, 18060-C-T, 28144-T-C), is the more remote ancestral haplotype. 3 cases from Washington collected around January 18 shared this 3-variant haplotype. The isolates were continuously present from late February through April 2020, with predominance found in the western states, including Washington and California, compared with isolates from the east coast. The potential immediate ancestral haplotypes inferred with CHLA CARD Genome Tracker were also from US and Canada isolates but they were sampled at later dates (Shen et al., 2020). This provided further evidence of reduced state-to-state and coast-to-coast transmissions within the United States. Based on the genomic analysis of all published SARS-CoV-2 sequences to date, safer at home measures have been very effective at reducing the spread of SARS-CoV-2. Persistent implementation of these measures would clearly lead to reduced spread of the COVID-19 pandemic over time. New haplotypes do not equate to new strains of the virus. The viral sequencing and analysis tools described here may prove to be critical, however, if a more transmissible and more deadly strain of SARS-CoV-2 emerges over time. Further studies will likely determine viral haplotypes, in the context of host factors, that may be associated with disease severity, response to treatment, or utility of vaccines for disease prevention.

## Data Availability

Data used in the manuscript were extracted from public data resources, namely GISAID (https://www.gisaid.org/) and GenBank (https://www.ncbi.nlm.nih.gov/genbank/sars-cov-2-seqs/). They can also be accessed at the Children's Hospital Los Angeles (CHLA) COVID-19 Analysis Research Database (CARD) (https://covid19.cpmbiodev.net/covid19/index.php).

